# Charted times of bedside pain assessments are slot labels: prevalence across hospitals, consequences for estimating analgesic onset, and when hourly charting is an adequate approximation

**DOI:** 10.64898/2026.11.14.26368116

**Authors:** Yangming Lee, Kris Moe

## Abstract

**Objective:** Temporal and causal analyses of electronic health records (EHRs) treat the charted time of an observation as the time it was made. We quantified how often bedside pain and sedation assessments are charted to a time slot, how this affects estimates of analgesic onset, and when hourly charting is an adequate approximation.

**Methods:** We used intensive care unit (ICU) data from one academic center (MIMIC-IV) and 78 hospitals (eICU-CRD). Slot labeling was identified from charted minutes and from the delay between charted and entry times. In MIMIC-IV, opioid administration times served as anchors, and faster onset of intravenous (IV) than oral opioids as a pharmacologic benchmark. We compared onset estimated from charted times with onset from exactly timed records, bounded true assessment times under alternative charting mechanisms, and used simulation to relate bias to the ratio of slot width to response time constant (Δ*/k*).

**Results:** In MIMIC-IV, 68% of pain and 76% of sedation scores were charted on the hour and entered a median of 41–44 minutes later. Of the first reassessments after a dose, 32–34% of on-hour records carried a time before the dose, against 12–14% of off-hour records. Across eICU hospitals, the on-hour share ranged from 15% to 92% (10th–90th percentile). By charted time, oral opioids appeared nearly as fast as IV (pain change at 15–30 minutes, *−*1.76 vs *−*2.11 points); exactly timed records showed *−*0.80 vs *−*2.00. Slower oral onset held under every charting mechanism compatible with the data, but its magnitude was not identified (ratio of time constants 1.2–11.6). In simulation, analysis by charted time was adequate up to Δ*/k ≈* 2 when rounding was to the nearest slot, but only up to Δ*/k ≈* 0.1 when rounding had a direction.

**Conclusion:** Charted times of bedside assessments are often slot labels, and charting practice is hospital-specific. For ICU analgesic onset, neither condition for an adequate approximation can be assumed, and EHR data alone cannot recover the size of the effect.

## 1 Introduction

Time series built from electronic health records (EHRs) are increasingly used for questions about order and timing: how fast a treatment acts, whether one event precedes another, and which variables drive which in time-series causal discovery, causal graph learning, and target-trial emulation [1–4]. These methods share an assumption that is rarely stated: the charted time of an observation is the time the observation was made.

For machine-generated data—such as continuous bedside physiological monitors, electroen-cephalography, or automated operating-room motion and video capture [5–7]—the assumption is close to true. For bedside assessments that a nurse makes and then charts—pain scores, sedation scores, neurological checks—it need not be. The nurse chooses which time to chart. A score obtained at 07:40 can be charted at 08:00 because that is the row of the flowsheet the nurse fills in, and entered at 08:35 when the nurse sits down to document. The charted time is then a *slot label*: it identifies the charting slot, not the moment of assessment. Delayed documentation and rounded times have been described for other EHR data (Section 2). What is missing is (i) a large, multicenter quantification for bedside assessments, (ii) a demonstration of what slot labels do to an effect estimate whose true value is known, and (iii) a rule for when charting to a slot is an acceptable approximation.

Pain and sedation scores are central variables in ICU care and research [8], and opioid onset gives a natural test case, because pharmacology fixes the answer qualitatively: IV opioids act within minutes, immediate-release oral opioids in about 40 minutes (Section 2.4). Any analysis of bedside pain scores that does not find IV opioids acting faster than oral opioids is measuring something other than pharmacology.

This study is a measurement and data-quality study. We asked four questions.

1. How many bedside assessments are charted to a slot, how long is the delay to entry, and how much does this vary between and within hospitals (MIMIC-IV and 78 eICU-CRD hospitals)?
2. How does analysis by charted time distort the estimated onset of opioids, judged against the benchmark that IV is faster than oral?
3. How much can be recovered from records whose times are exact, and from bounding the true assessment time with exactly timed doses under all charting mechanisms compatible with the data?
4. How does the bias of slot charting depend on the ratio of slot width to the response time constant (Δ*/k*) and on the direction of rounding (simulation)?

We do not claim to solve the problem. Exactly timed doses can bound the true time of an assessment, but where in that bound it lies depends on a charting mechanism that, as we show, the data do not identify. Corrections are therefore reported as sensitivity analyses under explicit assumptions.

## 2 Related Work

### 2.1 Timing of EHR documentation

EHR data reflect the processes of care that generate them [9, 10]. Delayed and batched documentation of vital signs [11, 12], timestamp errors and late documentation of perioperative events [13, 14], heaping of manually entered times on round minutes [15, 16], and gaps between documented and actual medication times [17, 18] have all been described. These studies show that recorded times can be wrong; they do not quantify slot labeling of bedside assessments across hospitals or show what it does to an effect estimate.

|  |  |
|---|---|
| **Problem or Issue** | Temporal and causal analyses of electronic health records assume that the charted time of an observation is the time it was made. For nurse-charted bedside assessments, this is often not true. |
| **What is Already Known** | Documentation is often delayed and follows care processes, and the timing of a measurement can itself be informative. How often bedside scores are charted to a time slot, and how this biases effect estimates, had not been quantified. |
| **What this Paper Adds** | In MIMIC-IV, 68% of pain and 76% of sedation scores were charted on the hour; across 78 eICU-CRD hospitals the share ranged from 15% to 92%. By charted time, oral opioids appeared nearly as fast as IV. A simulation-based rule (Δ/*k*) shows when hourly charting is an adequate approximation. |
| **Who would benefit** | Informaticians and clinical researchers who use charted assessments in time-series, causal or target-trial analyses, and teams that design clinical documentation. |

### 2.2 Informative observation timing and coarse data

When an observation is made can depend on the patient’s state, so observation timing in EHRs is itself informative [19–22]. A slot label is a coarsened time, and coarsening is ignorable only if it occurs at random [23, 24]. Related statistical tools include interval-censored data [25], measurement error [26], multiple imputation [27] and partial identification under weak assumptions [28]; we use multiple imputation to carry timing uncertainty into effect estimates, and partial identification to bound true assessment times.

### 2.3 Time resolution in temporal and causal inference

Time aggregation, discrete-time approximation of time-dependent dynamics, and sub-sampling are known to distort inferred causal links and system identification [29, 30]; lagged EHR associations can show diseases following their own effects because of workflow [31]; and misplaced time zero biases effect estimates [32], even when time zero is abstracted from records [33]. These results concern the resolution or anchoring of time in general, not slot labels whose direction of rounding is unknown.

### 2.4 Pain and sedation assessment and opioid onset

Pain and sedation scores are outcomes in ICU care and research, the basis of dosing decisions, and the content of reassessments after analgesic doses, which accreditation standards require to be documented [34]; systematic assessment has been associated with better outcomes [35, 36], although documented reassessment within an hour of an opioid dose can be uncommon [37]. Pharmacology fixes the expected order of opioid onset. IV hydromorphone relieved experimental pain in volunteers within 5 minutes [38], the effect-site equilibration half-life of fentanyl is about 6 minutes (EEG endpoint) [39], and after an IV morphine bolus in postoperative patients the effect-site concentration peaks at about 20 minutes [40]; immediate-release oral oxycodone, by contrast, had a median onset of 41 minutes after surgery [41].

## 3 Methods

### 3.1 Data

MIMIC-IV v3.1 contains de-identified records of patients admitted to the ICUs of Beth Israel Deaconess Medical Center, Boston, from 2008 to 2022 [42, 43]. We included the first ICU stay of adults with a stay of at least 24 hours (51,838 stays). eICU-CRD v2.0 contains ICU stays at 208 US hospitals discharged in 2014–2015 [44, 45]. Both databases are available on PhysioNet [46] under a data use agreement. Use of the de-identified data is covered by the source institutions’ review board determinations (waiver of informed consent for MIMIC-IV; exemption for eICU-CRD). All patient-level processing took place in the cloud database and on a secure institutional server; only aggregate results left that environment, and no reported cell contains fewer than 10 patients. We followed the RECORD reporting guideline where applicable [47].

### 3.2 Identifying slot labels

In MIMIC-IV every charted observation carries a charted time and an entry time (storage time). We analyzed numeric pain scores (0–10), the post-intervention pain reassessment item, the Critical-Care Pain Observation Tool (CPOT), the Richmond Agitation–Sedation Scale (RASS), and, as a machine-generated contrast, heart rate validated from the monitor. A record was *on the hour* if its charted minute was 0. If times were not rounded, 1/60 (1.7%) of records would fall on the hour. We summarized the entry delay (entry time minus charted time) separately for on-hour and off-hour records, including the share entered *before* the charted time, which is only possible if the record was charted to a later slot than the one in which it was entered. We also examined whether scores sharing a nurse, patient and charted hour were entered together (batch entry), and how the on-hour share changed by unit and calendar period.

In eICU-CRD, times are minute offsets from ICU admission; the clock minute is the admission clock time plus the offset, modulo 60. The nurse charting table stores both an observation offset and an entry offset. We classified the charted minute as :00, :30, :15 or :45, another multiple of 5, or other. We analyzed hospitals with at least 1,000 pain scores and 50 ICU stays (78 hospitals). In some hospitals the entry offset equaled the observation offset for nearly every record, so the entry time carries no information; we called a hospital’s entry time *uninformative* if at least 90% of its records had an entry delay of exactly 0, and analyzed entry delays only in the remaining hospitals.

### 3.3 Multicenter analysis

We decomposed the variance of the on-hour indicator into between-hospital, between-unit (within hospital), between-stay (within unit) and within-stay components using the law of total variance, weighting by record. We assessed stability by correlating hospital on-hour shares between random halves of ICU stays and between discharge years 2014 and 2015, and consistency across measures by correlating the on-hour shares of pain, sedation and heart rate within hospitals. We related the on-hour share to hospital characteristics (teaching status, bed count, region) and to the type of entry time by linear regression. Finally, we compared the mean pain score of on-hour and off-hour records within each hospital (Wilcoxon signed-rank test across hospitals).

### 3.4 Dose anchors and pharmacologic benchmark

In MIMIC-IV, oral opioid doses come from bar-code medication administration records, whose times are exact (2–8% on the hour, 78–96% entered within 5 minutes). IV opioid boluses come from the ICU input records (fentanyl, hydromorphone and morphine pushes). These times are themselves partly rounded (41% on a quarter hour, including 21% on the hour, in the analysis cohort); we therefore repeated the main analysis using only IV doses charted off the quarter hour and entered within 5 minutes (45% of IV doses).

The analysis cohort consisted of single opioid doses given when the last pain score in the preceding 2 hours was at least 4, with no other analgesic or sedative event between the dose and the outcome score (58,987 doses). Outcome scores were pain scores and reassessments entered after the dose and charted between 60 minutes before and 240 minutes after it; we included scores charted before the dose but entered after it, because these may be post-dose reassessments charted to an earlier slot. The pharmacologic benchmark was that IV opioids act faster than oral opioids [38–41].

### 3.5 Onset estimates

We classified outcome records into three kinds. *Exactly timed records* were charted off the hour and entered within 5 minutes of the charted time; for these we took the charted time as the assessment time, and excluded records whose time fell before the dose. *Slot records* were charted on the hour. Other off-hour records, entered more than 5 minutes after the charted time, were counted separately in descriptive summaries and excluded from the exactly timed onset curves. For each record we computed the change from the pre-dose score. Onset curves averaged the change within patient and time bin (0–15, 15–30, 30–45, 45–60, 60–90, 90–120, 120–180, 180–240 minutes) and then across patients, so that standard errors are clustered by ICU stay. We also fitted an exponential onset model, change = *−E* (1 *− e^−t/k^*) for *t ∈* [0, 90] minutes, where *k* is the onset time constant (the time to about 63% of the maximum effect), and report the ratio *k*_oral_*/k*_IV_ and the IV minus oral difference at 15–30 minutes. Intervals came from 1,000 cluster-bootstrap samples of ICU stays (500 for the exact-dose sensitivity analysis).

### 3.6 Bounding the true assessment time

For a slot record charted at *c* and entered at *s* after a dose at *t_d_*, the true assessment time *T* satisfies

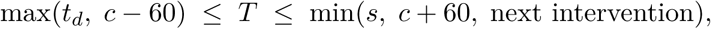

in minutes: it must follow the dose and precede entry and the next intervention, and a record charted to the hour slot is assumed to be no more than one slot away. The true time is thus interval-censored [25], or, in the terminology of coarse data, the charted time is a coarsening of it [23]. Where *T* lies within this interval depends on the charting mechanism. We modeled the mechanism as a mixture of rounding rules (to the previous, nearest or next hour) with a log-normal entry delay, fitted separately for IV and oral doses to the observed histogram of *s − c* among post-dose slot records. As a second moment, we used the share of first reassessments charted before the dose, after subtracting the share seen among exactly timed records (assessments that truly preceded the dose).

Because the mechanism was not identified (Results), we treated it as a sensitivity parameter. For each route we varied the shares of the three rounding rules over a simplex grid in steps of 0.25 (15 rules per route, 225 combinations), refitted the entry-delay distribution for each rule, and imputed true times by drawing from the posterior proportional to (prior *×* rounding rule *×* entry-delay density), analogous to posterior-based probabilistic data association and latent-state estimation under measurement uncertainty [48, 49], with the prior taken from the timing of exactly timed reassessments. We call a rule *compatible* if it fitted the *s − c* histogram about as well as the best rule (4 IV and 3 oral rules; 12 combinations). For the compatible combinations, we combined the cluster bootstrap with two imputations per bootstrap sample (250 samples) and estimated variances with the method of von Hippel and Bartlett [27]. This is an analysis of what the data allow under each assumption, not an estimate of the truth [28].

### 3.7 Representativeness of exactly timed records

Exactly timed records could differ from slot records in ways that matter for onset. We compared covariates of patients, doses and units (standardized mean differences), estimated how well patient and dose covariates predict whether a record is exactly timed (cross-validated area under the curve) compared with the identity of the charting nurse (leave-one-out), measured the split-half reliability of each nurse’s share of exactly timed records, compared the late plateau (120–240 minutes, where timing errors matter little) between the two kinds of record, and reweighted exactly timed records to the covariate distribution of all records (weights capped at the 99th percentile).

### 3.8 Simulation

#### Consistency check

To check the implementation of the imputation, we used exactly timed records as the truth, converted 72% of them into slot records with a known mechanism, and re-estimated onset (20 replicates). The same mechanism generated and corrected the data, so this tests only the code, not the assumptions (Supplementary Section A).

#### Time-scale simulation

To ask when slot charting is an acceptable approximation, we expressed time in units of the onset time constant (*k* = 1). True assessment times were uniform on [0, 6*k*], the change was *−*2(1 *− e^−T/k^*) plus normal noise (SD 2), and the dose fell at a uniform phase within the slot. Two processes with *k* = 1 and *k* = 2 (true ratio 2) each had 3,000 assessments; we varied Δ*/k* from 0.05 to 6 and the rounding rule (previous, nearest, next slot), with 200 replicates. We compared analysis by charted time, the slot midpoint, imputation with the correct mechanism, and imputation with a misspecified mechanism (for example, assuming nearest-slot rounding when rounding was to the previous slot). Adequacy was defined as a relative bias in *k* within *±*10%.

## 4 Results

### 4.1 Slot labels in MIMIC-IV

Bedside assessments were mostly charted on the hour: 68% of pain scores, 71% of pain reassessments, 78% of CPOT scores and 76% of RASS scores, against 1.7% expected without rounding (Table 1). On-hour and off-hour records were charted differently (Fig. 1). Off-hour records were entered almost at once: median delay 0–1 minute, 71–80% within 5 minutes, and fewer than 1% entered before their charted time. On-hour records were entered a median of 41–44 minutes after their charted time, 38–41% more than an hour later, and 15–17% *before* their charted time—that is, they were charted to a later hour than the one in which they were entered. Heart rate, whose time is set by the monitor, was also mostly charted on the hour (hourly validation) but was entered before its charted time in only 0.9% of records.

**Figure 1:**
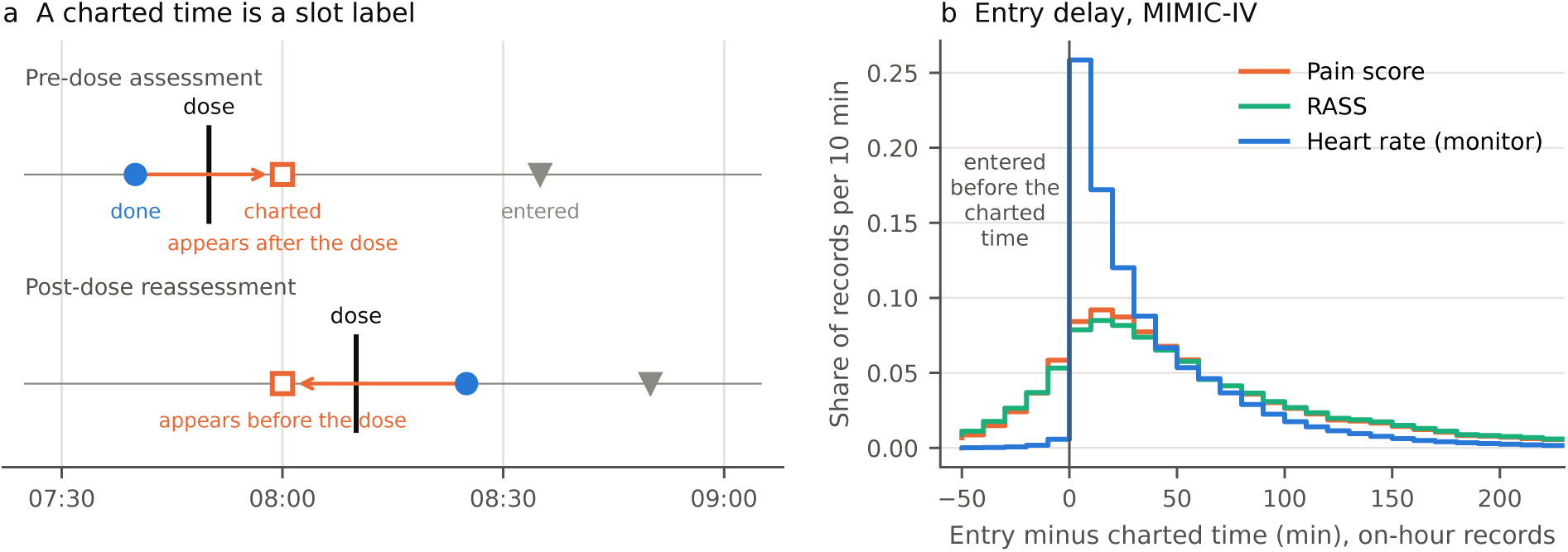
Slot labels in MIMIC-IV. **a**, Schematic. A score obtained at 07:40, before a dose at 07:50, is charted in the 08:00 slot and so appears to follow the dose; a reassessment made at 08:25, after a dose at 08:10, is charted at 08:00 and appears to precede it. **b**, Entry time minus charted time for records charted on the hour. Pain and RASS scores are often entered before their charted time (charted to a later hour); heart rate, timed by the monitor, almost never is. The last bin (*≥*230 minutes) is not shown.

**Table 1:** Slot labels and entry delays.

|  |  | Entry delay of on-hour records |  |  |  |
|---|---|---|---|---|---|
| Measure (MIMIC-IV) | On the hour | Median (min) | Before charted | >60 min | Off-hour: ≤5 min<sup>a</sup> |
| Pain score (0–10) | 68.4% | 41 | 14.9% | 37.8% | 72.9% |
| Pain reassessment | 71.3% | 44 | 15.1% | 40.4% | 70.7% |
| CPOT | 77.7% | 44 | 17.1% | 40.8% | 74.0% |
| RASS | 75.7% | 43 | 15.4% | 39.9% | 80.0% |
| Heart rate (monitor) | 87.4% | 24 | 0.9% | 22.8% | 26.3% |
| eICU-CRD, pain scores, 78 hospitals: median [10th, 90th percentile] across hospitals |  |  |  |  |  |
| Share charted at :00 |  | 37% [15%, 92%] (pooled 54.5%; expected 1.7%) |  |  |  |
| Entry time uninformative<sup>b</sup> |  | 48 of 78 hospitals |  |  |  |
| Median entry delay, :00 records<sup>c</sup> |  | 61 min (30 hospitals with informative entry time) |  |  |  |
| 90th percentile of entry delay, :00 records<sup>c</sup> |  | 216 min |  |  |  |
| :00 records entered >60 min late<sup>c</sup> |  | 50% |  |  |  |

<sup>a</sup> Share of off-hour records entered within 5 minutes of the charted time.
<sup>b</sup> At least 90% of records with entry offset equal to observation offset.
<sup>c</sup> Median across the 30 hospitals whose entry time is informative.

Pain and sedation scores were entered in batches: when a nurse charted both for the same patient at the same hour, 89% of pairs were entered within one minute of each other. The on-hour share was higher in the 2020–2022 anchor-year group than in 2008–2010 in every unit (for example, from 64% in 2008–2010 to 87% in 2020–2022 in the trauma surgical ICU, and from 63% to 82% in the medical ICU), and the median entry delay lengthened (medical ICU, 17 to 41 minutes).

The exact bar-code dose times show the consequence for ordering. Among all first reassessments entered after a dose, 32.2% of those after bar-code doses carried a charted time *before* the dose (26.8% after IV pushes). Some reassessments do truly precede a dose (for example, a reassessment of the previous dose entered late): among off-hour first reassessments in the analysis cohort, this share was 11.9% (oral) to 13.7% (IV). Among on-hour first reassessments it was 33.9% (oral) and 32.0% (IV), so roughly 20 percentage points can be attributed to slot charting.

### 4.2 Multicenter: 78 eICU hospitals

The 78 hospitals contributed 2.41 million pain scores. Pooled, 54.5% were charted at :00, with further heaping at :30, :15 and :45 and at other multiples of 5 minutes (Fig. 2a). The on-hour share varied widely between hospitals: 10th percentile 15%, median 37%, 90th percentile 92%. In 48 hospitals the entry offset equaled the observation offset for essentially every record (median 100%), so the entry time carries no information, possibly because those records were imported from the hospital’s own EHR. In the other 30 hospitals the median on-hour share was 86% (25% in the 48), on-hour records were entered a median of 61 minutes late (median across hospitals; 90th percentile 216 minutes), and half were entered more than an hour late.

**Figure 2:**
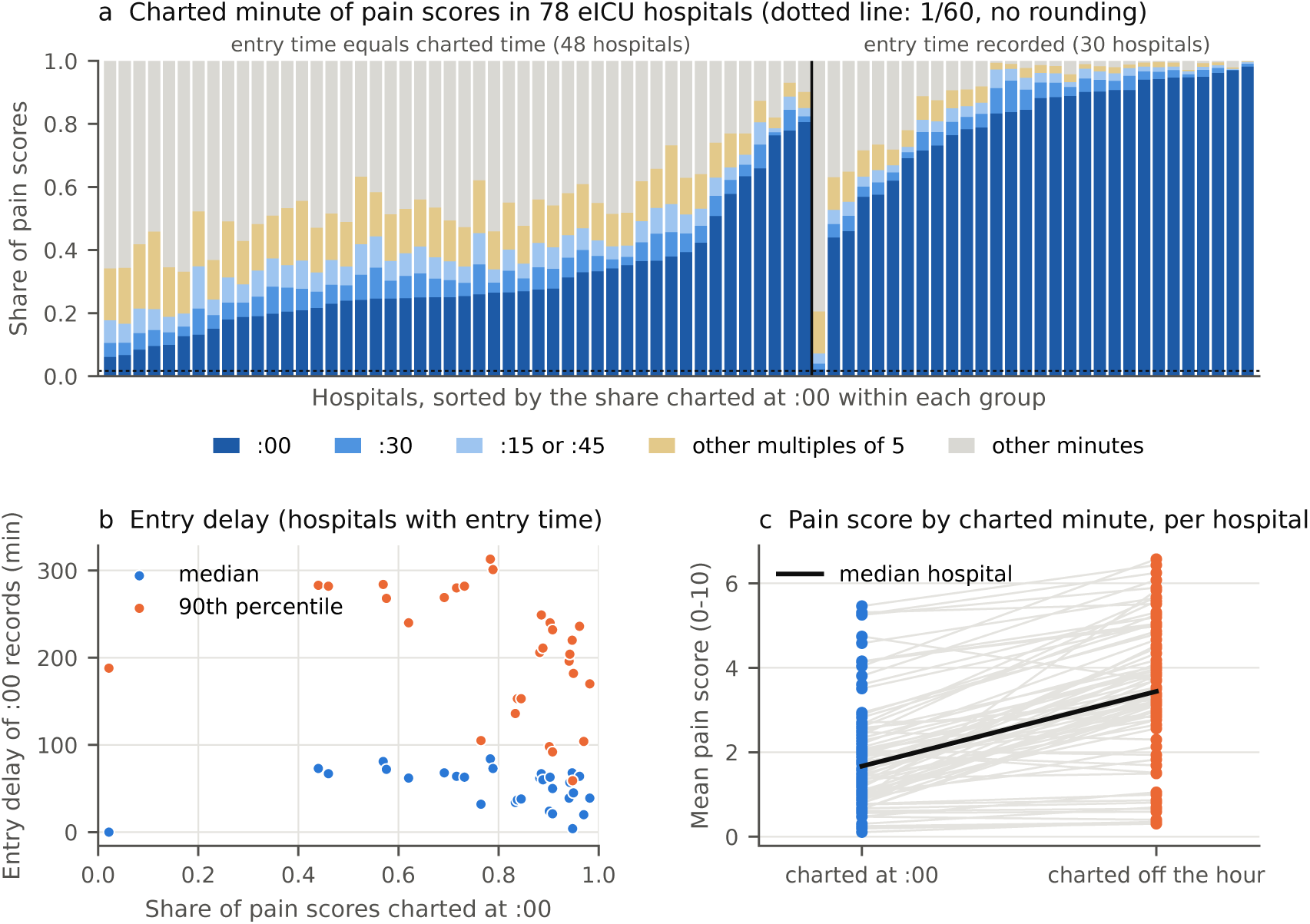
Slot labels across 78 eICU-CRD hospitals. **a**, Charted clock minute of pain scores by hospital; hospitals are grouped by whether the entry time is informative and sorted by the share charted at :00. The dotted line marks 1/60, the share expected at :00 without rounding. **b**, Median and 90th percentile of the entry delay of :00 records against the share charted at :00, in the 30 hospitals with informative entry times. **c**, Mean pain score of records charted at :00 and off the hour in each hospital (lines join the same hospital).

Slot labeling was a property of the hospital, not the unit. Between-hospital variance accounted for 28% of the variance of the on-hour indicator, between-unit variance within hospitals for 0.6%, between-stay variance for 12% and within-stay variance for 59% (sedation scores: 23%, 1.7%, 11% and 65%). Hospital on-hour shares were almost identical between random halves of stays (*r* = 0.999) and between 2014 and 2015 (*r* = 0.97), and correlated across measures within a hospital (pain vs sedation *r* = 0.78; pain vs heart rate *r* = 0.85), consistent with a hospital-wide flowsheet configuration. Teaching status, bed count and region together explained little (adjusted *R*^2^ = 0.19); the type of entry time alone explained more (adjusted *R*^2^ = 0.60). Among the 30 hospitals with informative entry times, a higher on-hour share went with *shorter* entry delays (Spearman *ρ* = *−*0.39); pooled over all 78 hospitals the correlation reversed (*ρ* = 0.72) because the two record systems differ in both (Fig. 2b).

Within hospitals, the timing of a record carried information about the patient (Fig. 2c). Off-hour pain scores were on average 1.96 points higher than on-hour scores in the same hospital (pooled means 3.80 vs 1.27), and the share of scores *≥* 4 was 27 percentage points higher; 66 of 71 hospitals with enough scores showed the difference (*p <* 10*^−^*^11^), in both kinds of record system. In MIMIC-IV the within-stay difference was smaller (0.51 points). MIMIC-IV has a 4-hourly scheduled assessment: at 00, 04, 08, 12, 16 and 20 h there were 5–10 times as many pain scores as at other hours, with means of 2.4–2.9 against 4.0–4.6. On-hour records thus mix routine scheduled assessments with event-triggered assessments charted to the hour.

### 4.3 Consequences for estimated onset

Analysis by charted time erased most of the difference between routes (Fig. 3a). At 15–30 minutes after the dose, pain had fallen by 2.11 points after IV and 1.76 points after oral opioids, and the fitted onset time constants were 7.2 minutes (IV) and 15.1 minutes (oral). An oral onset of 15 minutes is pharmacologically implausible [41]; it arises because charted-time bins mix in low-pain routine scheduled :00 assessments and post-dose reassessments rounded backward to the :00 slot just after the dose, producing a steep apparent early drop followed by an upward rebound after 60 minutes. In exactly timed records (5,538 ICU stays), pain at 15–30 minutes had fallen by 2.00 points after IV opioids but only 0.80 points after oral opioids, and oral opioids reached the IV effect only at 45–60 minutes (Fig. 3b; the slight positive change at 0–15 minutes after oral doses reflects pain worsening between the last pre-dose score in the preceding 2-hour window and actual administration, before oral absorption begins). The fitted time constant was 14.9 minutes for IV opioids [95% cluster-bootstrap interval 12.3, 18.1] and 80 minutes for oral opioids [55, 132], a ratio of 5.4 [3.5, 9.3]; the IV minus oral difference at 15–30 minutes was *−*1.19 [*−*1.57, *−*0.83] (Table 2). The oral time constant is poorly determined because the oral curve had not reached its plateau by 90 minutes; the direction and the early difference are the robust quantities.

**Figure 3:**
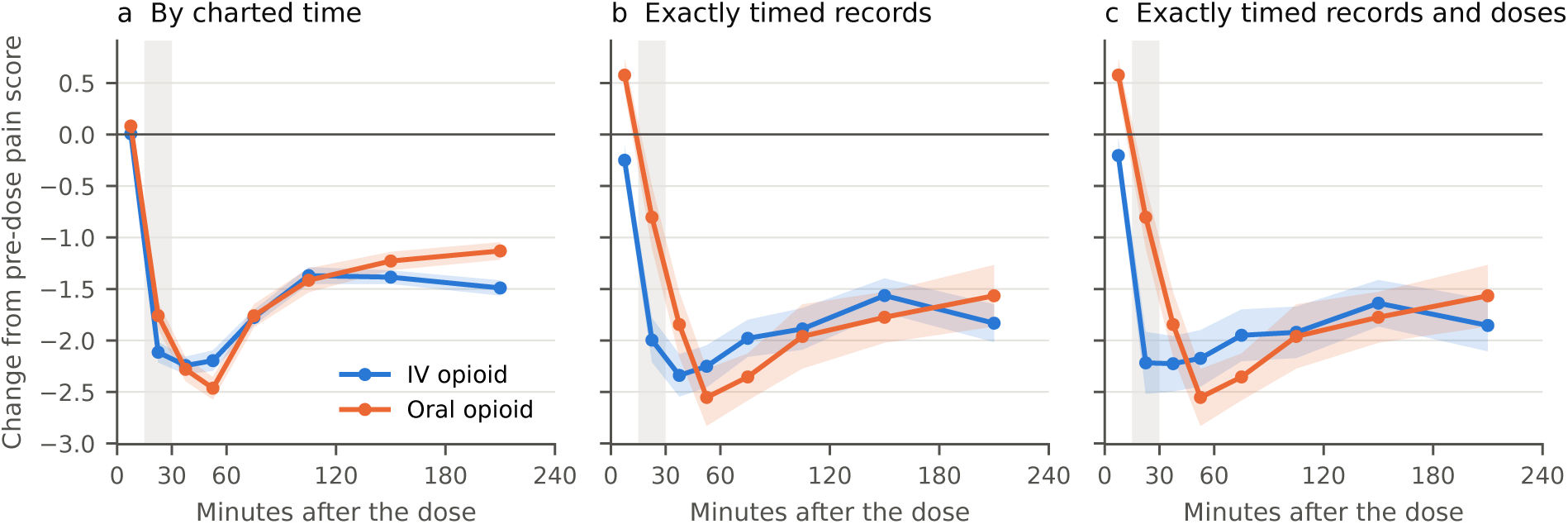
Change in pain score after single opioid doses in MIMIC-IV, by the time used for the assessment. **a**, Charted time (all records). **b**, Exactly timed records only (charted off the hour and entered within 5 minutes). **c**, Exactly timed records after exactly timed IV doses only (off the quarter hour and entered within 5 minutes); oral doses are bar-code scans in all panels. Points are means across ICU stays of within-stay means; bands are 95% confidence intervals clustered by stay. The gray band marks 15–30 minutes.

**Table 2:** Onset estimates under different treatments of time (MIMIC-IV).

| Analysis | *k*<sub>IV</sub> (min) | *k*<sub>oral</sub> (min) | *k*<sub>oral</sub>/*k*<sub>IV</sub> | IV − oral, 15–30 min |
|---|---|---|---|---|
| Charted time | 7.2 | 15.1 | 2.1 | −0.35 |
| Entry time (upper bound) | 33.8 | 300<sup>a</sup> | 8.9 | — |
| Exactly timed records | 14.9 [12.3, 18.1] | 80 [55, 132] | 5.4 [3.5, 9.3] | −1.19 [−1.57, −0.83] |
| exactly timed IV doses only | 11.7 [9.3, 14.6] | 80 [56, 137] | 6.9 [4.5, 12.1] | −1.41 [−1.85, −1.01] |
| reweighted to all records | 17.5 [14.7, 21.8] | 127 [72, 154] | 7.2 [4.1, 9.4] | −1.26 [−1.67, −0.83] |
| Imputation, 12 compatible mechanisms<sup>b</sup> | 6.0–26.7 | 31.6–70.7 | 1.2–11.6 | — |
| union of intervals | 4.8–29.2 | 28.9–83.5 | 1.05–15.3 | — |

Brackets: 95% cluster-bootstrap intervals (ICU stays). *k* is the time constant of −*E*(1 − *e*<sup>−*t*/*k*</sup>) fitted over 0–90 minutes.
<sup>a</sup> At the upper bound of the fit.
<sup>b</sup> Range of point estimates over the 12 combinations of IV and oral charting rules compatible with the entry-delay histogram; all 12 intervals for the ratio exclude 1.

Restricting to exactly timed IV doses (8,577 IV and 3,189 oral records) made the IV onset faster: *k*_IV_ = 11.7 minutes [9.3, 14.6], ratio 6.9 [4.5, 12.1], difference at 15–30 minutes *−*1.41 [*−*1.85, *−*1.01] (Fig. 3c). Rounded dose times had blurred the IV curve in the same way as rounded assessment times: error in the time variable attenuates the estimated response, as measurement error in a covariate does [26, 50].

Using the entry time instead of the charted time over-corrected (IV 33.8 minutes; the oral fit reached the upper bound), and the slot midpoint gave 35.3 and 91.2 minutes. Neither is a defensible estimate: the entry time is an upper bound on the assessment time, and the midpoint assumes symmetric rounding.

### 4.4 How much can be recovered

#### The charting mechanism is not identified

Fitted to the entry-delay histogram, post-dose slot records were best described by mostly nearest-hour rounding (IV 90% nearest and 10% to the previous hour, median entry delay 40 minutes; oral 90% nearest and 10% to the next hour, median 60 minutes). The histogram constrained the entry-delay median well but did not separate rounding rules: 4 IV and 3 oral rules fitted about equally well. The second moment, the share of first reassessments charted before the dose, should separate them, since rounding to the previous hour moves records before the dose. After subtracting the share of assessments that truly preceded the dose, the targets were 21% (IV) and 25% (oral). No rounding rule matched both moments: rules compatible with the histogram predicted at most 15%, and only rules dominated by rounding to the previous hour reached the target, at the cost of a poor histogram fit. Simple rounding of the true assessment time to a slot is therefore rejected by the data; other practices, such as charting the reassessment in the row of the dose or charting the time at which a reassessment was due, would be needed and are not identifiable from these records.

#### Direction robust, magnitude not identified

Across all 225 combinations of IV and oral rules, the ratio *k*_oral_*/k*_IV_ ranged from 0.45 to 32.6 (Supplementary Fig. S1). Ratios below 1 arose only when IV records were assumed to be rounded mostly to the previous hour and oral records mostly to the nearest or next hour, a pairing that contradicts the histogram for both routes. Under each of the 12 compatible combinations, oral onset was slower (Fig. 4): the ratio ranged from 1.2 to 11.6 and every 95% interval excluded 1 (union of intervals 1.05–15.3). The IV time constant ranged from 6 to 27 minutes and the oral one from 32 to 71 minutes, depending on the assumed mechanism.

**Figure 4:**
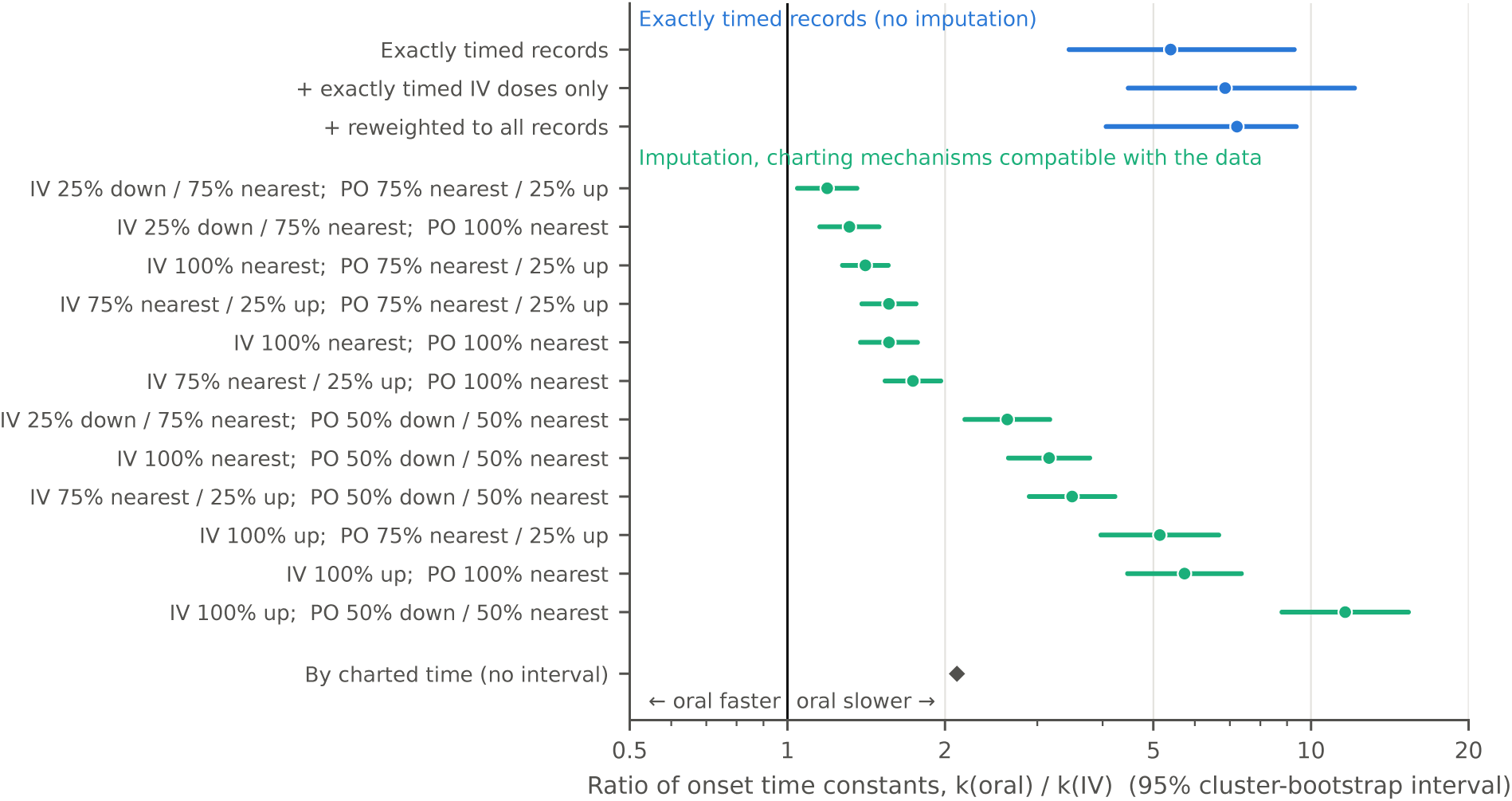
Ratio of oral to IV onset time constants under each analysis. Blue: exactly timed records, with the exact-dose and reweighting sensitivity analyses. Green: imputation of true assessment times under each of the 12 combinations of charting rules compatible with the entry-delay histogram (rules: share rounded to the previous hour / nearest hour / next hour). Lines: 95% cluster-bootstrap intervals (von Hippel–Bartlett variance for imputation). Diamond: charted time.

#### Exactly timed records are informative but not a random sample

Exactly timed and slot records were similar in covariates (all standardized mean differences below 0.24). Whether a record was exactly timed was predicted poorly by patient and dose covariates (cross-validated AUC 0.61) and well by the charting nurse (AUC 0.84); nurses’ shares of exactly timed records ranged from 0.3% to 35% (10th–90th percentile) and were stable (split-half *r* = 0.91, 386 nurses). Charting habit, rather than the patient’s condition, thus largely decides which records are exactly timed. However, at the late plateau, where timing errors matter little, exactly timed records showed slightly larger reductions than slot records (IV *−*0.17 [*−*0.31, *−*0.04]; oral *−*0.52 [*−*0.70, *−*0.31] points). Reweighting exactly timed records to the covariate distribution of all records left the conclusions unchanged (Table 2).

### 4.5 When is slot charting an adequate approximation?

In the time-scale simulation, the direction of rounding mattered more than the width of the slot (Fig. 5). With rounding to the nearest slot, the error in assessment time is symmetric, and analysis by charted time estimated *k* within 10% up to Δ*/k* = 2. With rounding to the previous or next slot the error has a direction, and analysis by charted time was within 10% only up to Δ*/k* = 0.1; at Δ*/k* = 0.25 the bias was already *−*16% (previous) or +17% (next), and at Δ*/k* = 1 it was *−*49% or +70%. The ratio of two processes was distorted accordingly (at Δ*/k* = 1, 2.8 or 1.6 instead of 2). Imputation with the correct mechanism was unbiased up to Δ*/k* = 2 but failed beyond 3, where the slot is too wide to retain the information; imputation with a misspecified mechanism was no better than the charted time, and the slot midpoint was within 10% up to Δ*/k* = 1.5.

**Figure 5:**
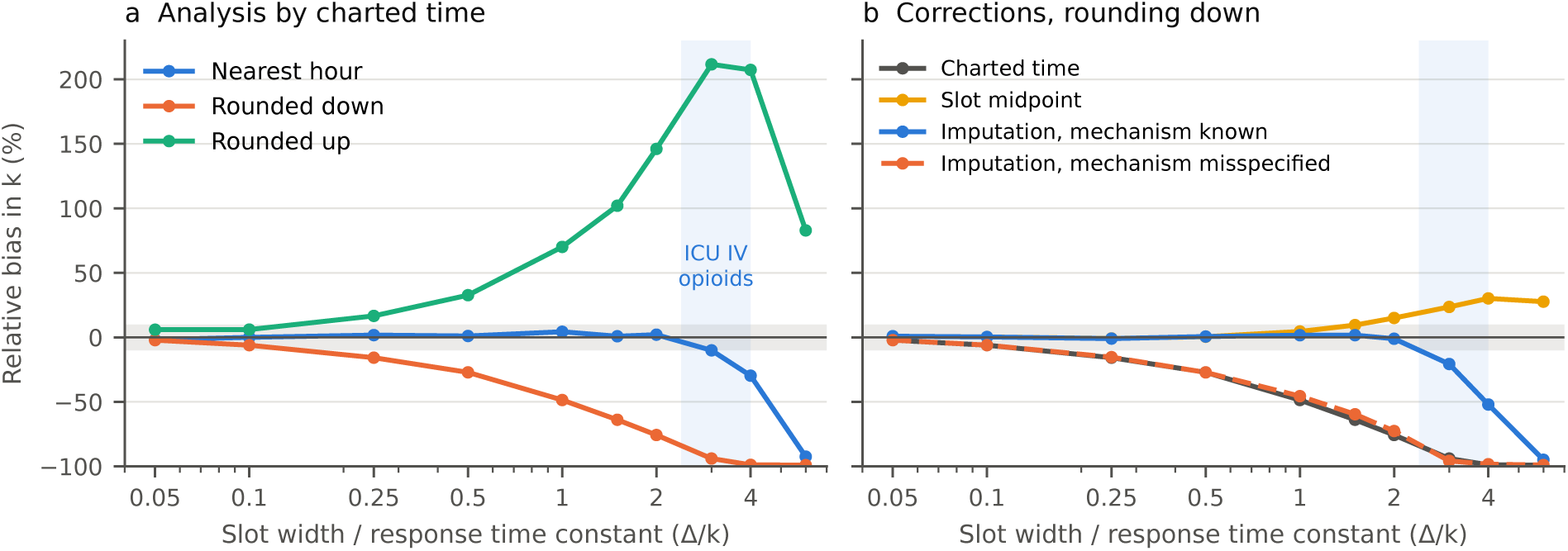
Relative bias of the estimated onset time constant *k* as a function of the ratio of slot width to time constant (Δ*/k*), in simulation (200 replicates). **a**, Analysis by charted time under three rounding rules. **b**, Corrections when rounding is to the previous slot: charted time, slot midpoint, imputation with the correct mechanism, and imputation assuming nearest-slot rounding. Gray band: *±*10%. Blue band: Δ*/k* of ICU IV opioids with hourly slots.

With hourly slots, ICU IV opioids (*k ≈* 12–25 minutes) have Δ*/k ≈* 2.4–5, at or beyond the limit even with a known mechanism, and oral opioids (*k ≈* 35–80 minutes) have Δ*/k ≈* 0.75–1.7, where the result depends on the direction of rounding—which, in MIMIC-IV, is exactly what the data do not identify.

## 5 Discussion

### 5.1 Principal findings

Charted times of bedside pain and sedation assessments are mostly slot labels. In MIMIC-IV, two thirds to three quarters of scores sit on the hour and are entered about 40 minutes later, and some are entered before the hour they are charted to. Across 78 eICU hospitals, the on-hour share ranges from about 15% to over 90%, is a stable property of the hospital rather than the unit, and in most hospitals cannot even be checked against an entry time, because the stored entry time is a copy of the charted time. Slot labels scramble order: about one in five post-dose reassessments charted on the hour is placed before the dose it follows. Analysis by charted time makes oral opioids look almost as fast as IV opioids. Records whose time is exact restore the pharmacology, but the size of the difference cannot be recovered from the rest of the data, because the charting mechanism is not identified. Whether charting to a slot is harmless depends on Δ*/k* and on the direction of rounding.

### 5.2 Implications for temporal and causal analysis

Methods that infer effects or causal structure from the order of events—lagged regression, Granger-type tests, time-series causal discovery, target-trial emulation with time-varying treatments—take the charted order as the true order. Coarse time resolution and misplaced time zero are known to distort such analyses (Section 2). Slot labels violate the assumed order exactly where the order matters most: around interventions, when nurses chart the assessment that prompted a dose and the reassessment that followed it in the same documentation session. In our data, a reassessment of a dose can appear before the dose, and an assessment that prompted a dose can appear after it. An analysis that conditions on “the last score before the dose” or “the first score after the dose” can therefore pick up the wrong score. The same applies to any subjective, nurse-charted variable (sedation, delirium screening, neurological checks); it does not apply to monitor-timed signals, whose charted times were essentially never later than their entry.

### 5.3 The timing of an assessment is informative

On-hour and off-hour scores differ by about 2 points within eICU hospitals. This is not an artifact to be averaged away: on-hour records are dominated by scheduled routine assessments, and off-hour records by event-triggered ones (a patient’s complaint, a reassessment after a dose). When an assessment is made, and therefore which time it is charted to, depends on the patient’s state. As for other EHR observations (Section 2), the degree of coarsening (exact time or slot label) and the timing itself depend on the patient’s state, so the coarsening is not at random: analyses that treat observation times as ignorable will be biased, and exactly timed subsets are not random samples of assessments. This also means that the measurement schedule is itself a clinical decision whose effect on treatment can be studied, which we address in separate work.

### 5.4 A checklist for analysts

1. Estimate Δ*/k*: the charting resolution relative to the time constant of the process studied. If Δ*/k ≤* 0.1, slot charting is harmless whatever the rounding.
2. Check whether the entry time is informative (in eICU, it is not in most hospitals) before using it to bound or classify records.
3. Estimate the on-hour share and entry delay per hospital, and analyze hospitals separately or adjust for them.
4. If 0.1 *<* Δ*/k ≤* 2, the result depends on the direction of rounding; report the assumed convention and analyze sensitivity to it. Imputation helps only if the mechanism is right.
5. If Δ*/k >* 2, use exactly timed subsets or better data, and report how representative those subsets are.
6. Check the anchors too: in our data, ICU bolus times were themselves partly rounded.

### 5.5 Implications for documentation

EHRs should distinguish the assessment time from the documentation time and should not pre-fill the assessment time with the flowsheet slot. Multicenter studies should report each site’s charting practice, including whether the entry time is real. Automated time capture and real-time clinical sensor or workflow logging avoid the heaping of manual entry [15, 51, 52]. Events with machine-recorded times (bar-code administration, infusion pumps, monitor alarms, device data) can serve as anchors, but only if their own times are exact [18].

### 5.6 Limitations

Dose anchors were available in one center only; eICU-CRD lacks consistently timed bar-code administration and entry records across hospitals, so the consequences for onset could not be examined across sites. The charting mechanism was not identified: the family of simple rounding rules was rejected by the data, and we did not model extended mechanisms (charting a reassessment in the dose’s row, or charting the time at which it was due). The semi-synthetic consistency check recovered *k* but did not recover the 15–30 minute difference (coverage of binned curves about 40%), and because it coarsened only true post-dose reassessments, it yielded a slower charted-time estimate rather than reproducing the artificially fast onset seen in the real data (where post-dose windows also include low-pain routine scheduled :00 assessments and reassessments charted when pain has improved). Exactly timed records are determined mainly by nurse habit but showed a slightly larger late effect. The oral time constant was poorly determined within 90 minutes. IV agents (fentanyl, hydromorphone, morphine) differ in onset by minutes, below the resolution of reassessment intervals, and could not be distinguished. The time-scale simulation is a simplified setting. Our data concern acute pain in ICUs; other settings have other slot widths (for example, daily headache diaries or visit-level ratings), and Δ*/k* must be estimated for each.

## 6 Conclusion

Charted times of bedside assessments are often slot labels, and charting practice differs between hospitals. Charting to a slot is an adequate approximation when Δ*/k* is small (about 0.1 or less) or when rounding is symmetric and Δ*/k* is at most about 2. For ICU analgesic onset neither condition can be assumed, and EHR data alone cannot recover the size of the effect. Recording the true assessment time is the remedy.

## Data Availability

All data produced are available online at:
https://physionet.org/content/mimiciv/3.1/
https://physionet.org/content/eicu-crd/2.0/

## Data and code availability

MIMIC-IV and eICU-CRD are available to credentialed users on PhysioNet. Analysis code and all aggregate results (with no cell containing fewer than 10 patients) are available from the authors upon reasonable request and will be deposited in a public repository upon publication.

## Ethics statement

This study is a secondary analysis of two de-identified public databases. The collection of patient information and the creation of MIMIC-IV were reviewed by the Institutional Review Board of Beth Israel Deaconess Medical Center, which granted a waiver of informed consent and approved the data sharing initiative. eICU-CRD is exempt from Institutional Review Board approval because of its retrospective design, the lack of direct patient intervention and its security schema, for which the re-identification risk was certified as meeting safe harbor standards by an independent privacy expert. The authors accessed both databases under PhysioNet credentialed access and data use agreements. Because only de-identified data were used, no additional ethics approval was required.

## CRediT authorship contribution statement

**Yangming Lee:** Conceptualization, Methodology, Software, Formal analysis, Writing – original draft. **Kris Moe:** Validation, Writing – review and editing.

## Acknowledgments

The authors thank the investigators and staff of the MIT Laboratory for Computational Physiology and Beth Israel Deaconess Medical Center for making MIMIC-IV and eICU-CRD available through PhysioNet.

## Declaration of generative AI and AI-assisted technologies in the manuscript preparation process

During the preparation of this work the authors used Google Gemini to polish the writing and proof reading. After using this tool, the authors reviewed and edited the content as needed and take full responsibility for the content of the published article.

## Supplementary material

### A Consistency check of the imputation

Exactly timed records served as the truth (*k*_IV_ = 14.9, *k*_oral_ = 80.5, ratio 5.4). In each of 20 replicates, 72% of them were converted to slot records under the route-specific mechanisms fitted in the real data (IV: 90% nearest, 10% previous hour, median entry delay 40 minutes; oral: 90% nearest, 10% next hour, median 60 minutes), and onset was re-estimated (Table S1). Imputation under the route-specific mechanism recovered *k*_IV_ and the ratio approximately, with interval coverage of 100% for *k*; analysis by charted time recovered *k*_IV_ but overestimated *k*_oral_ and the ratio; the entry time and the midpoint overestimated both. When both routes were converted with a single shared mechanism, every method was biased for *k*_oral_. In an earlier version that used a single mechanism for both routes, imputation assuming uniform timing within the interval was biased (IV *k* 8.8, coverage 0%). Binned onset curves were flattened by every method (15–30 minute difference: truth *−*1.19; methods *−*0.74 to *−*1.16; coverage about 40%). Because the same mechanism generated and corrected the data, this checks the implementation only.

**Table S1:** Consistency check (route-specific mechanisms; mean over 20 replicates).

| Method | *k*<sub>IV</sub> | *k*<sub>oral</sub> | Ratio |
|---|---|---|---|
| Truth (exactly timed records) | 14.9 | 80.5 | 5.4 |
| Charted time | 15.0 | 129.0 | 8.6 |
| Entry time | 27.1 | 269.4 | 10.1 |
| Slot midpoint | 25.0 | 163.9 | 6.6 |
| Imputation, pooled mechanism | 15.0 | 158.4 | 10.6 |
| Imputation, route-specific mechanism | 14.4 | 91.3 | 6.3 |

### B All 225 charting-rule combinations

### C eICU-CRD details

**Table S2:**
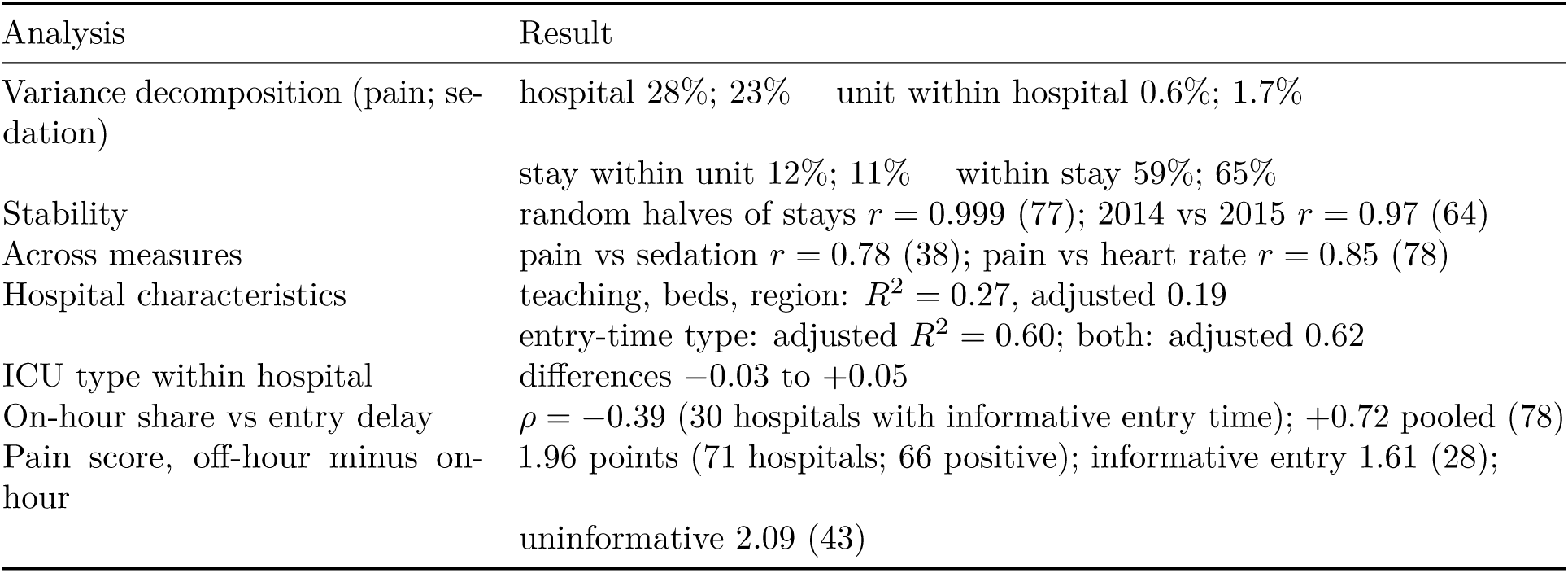
Hospital-level analyses of the on-hour share of pain scores (78 hospitals).

| Analysis | Result |
|---|---|
| Variance decomposition (pain; se-dation) | hospital 28%; 23% unit within hospital 0.6%; 1.7% |
|  | stay within unit 12%; 11% within stay 59%; 65% |
| Stability | random halves of stays *r* = 0.999 (77); 2014 vs 2015 *r* = 0.97 (64) |
| Across measures | pain vs sedation *r* = 0.78 (38); pain vs heart rate *r* = 0.85 (78) |
| Hospital characteristics | teaching, beds, region: *R*<sup>2</sup> = 0.27, adjusted 0.19 |
|  | entry-time type: adjusted *R*<sup>2</sup> = 0.60; both: adjusted 0.62 |
| ICU type within hospital | differences −0.03 to +0.05 |
| On-hour share vs entry delay | *ρ* = −0.39 (30 hospitals with informative entry time); +0.72 pooled (78) |
| Pain score, off-hour minus on-hour | 1.96 points (71 hospitals; 66 positive); informative entry 1.61 (28); |
|  | uninformative 2.09 (43) |

**Figure S1:**
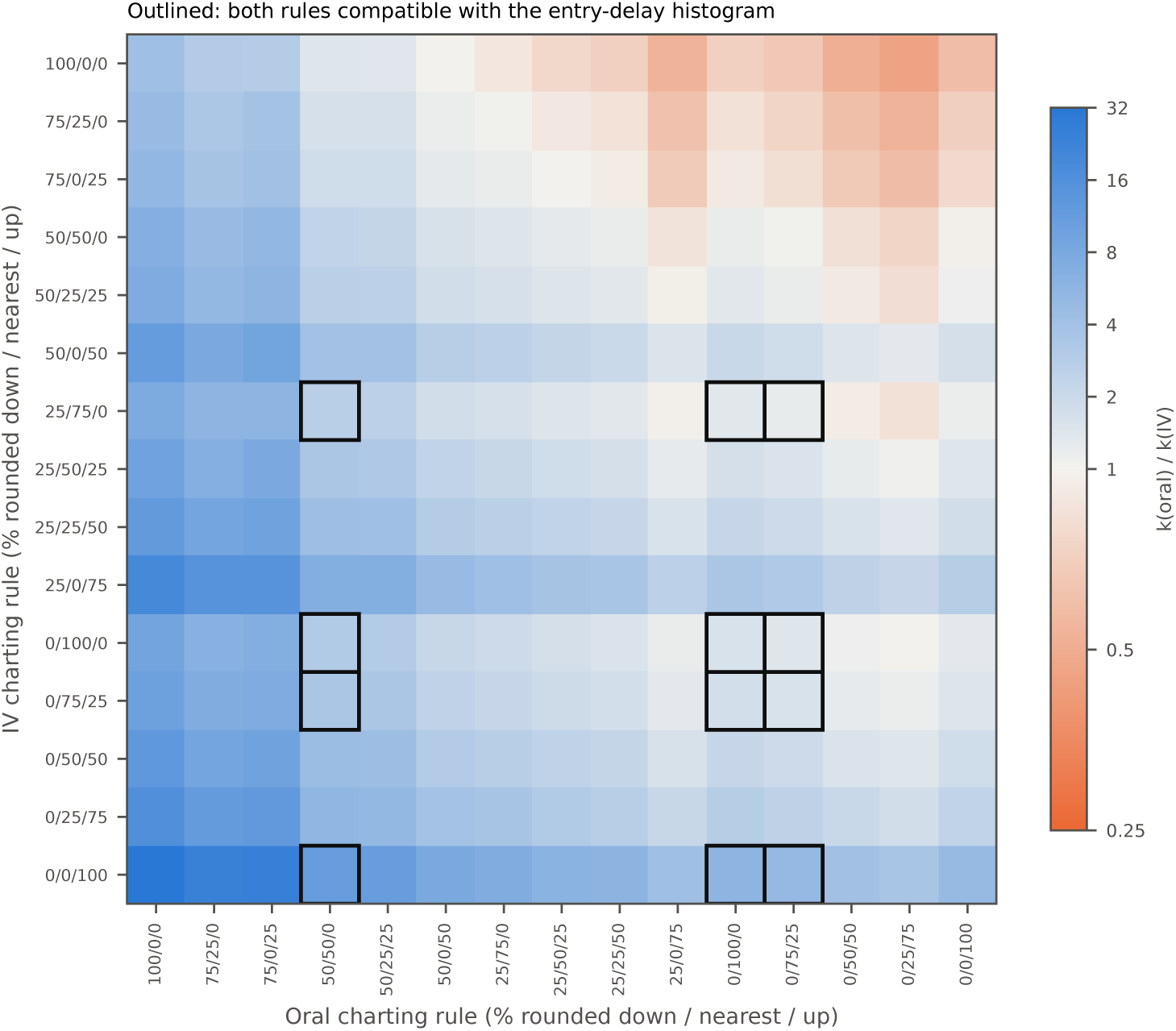
Ratio *k*_oral_*/k*_IV_ (point estimate) under each combination of IV and oral charting rules. Each rule is the share of slot records rounded to the previous hour / nearest hour / next hour; the entry-delay distribution was refitted for each rule. Outlined cells: both rules compatible with the entry-delay histogram. Of the 225 combinations, 173 had intervals entirely above 1, 34 included 1 and 18 were below 1.

